# SurvTEDVAE: A Disentangled Variational Autoencoder for Heterogeneous Treatment Effect Estimation with Time-to-Event Outcomes

**DOI:** 10.64898/2026.04.26.26351790

**Authors:** William JB Powell, Linying Zhang

## Abstract

Estimating heterogeneous treatment effects (HTE) from observational health data is essential for precision medicine, yet existing methods often struggle with high-dimensional covariates and time-to-event outcomes common in electronic health records (EHRs). We propose SurvTEDVAE, a disentangled variational autoencoder designed for causal survival analysis. The model learns latent representations corresponding to instrumental factors, confounders, and outcome-dependent risk factors, and integrates a survival likelihood to model time-to-event outcomes with censoring. The learned representations are used to estimate conditional average treatment effects using downstream causal estimators. We evaluated SurvTEDVAE using a semi-synthetic ACTG dataset and a high-dimensional EHR-based hypertension cohort with over 20,000 covariates. Across both datasets, SurvTEDVAE achieved lower estimation error for heterogeneous treatment effects compared with meta-learning and causal survival forest approaches. These results demonstrate that disentangled representation learning can improve causal effect estimation for survival outcomes in high-dimensional real-world health data.

## 1 Introduction

Real-world data, such as electronic health records (EHRs), have transformed clinical research by enabling large-scale evaluation of treatment effectiveness and safety in real-world populations. Regulatory agencies, including the U.S. Food and Drug Administration (FDA) and the European Medicines Agency (EMA), increasingly rely on real-world evidence (RWE) for post-market safety surveillance and comparative effectiveness research in diverse and heterogeneous real-world populations [1, 2, 3].

While randomized controlled trials (RCTs) remain the gold standard for drug efficacy and safety, their restrictive eligibility criteria and relatively small sample size often limit generalizability of their findings and reduce power to detect treatment heterogeneity across subpopulations[4]. In contrast, real-world data capture broader populations with substantial variability in demographics, comorbidities, and care pathways. Its relatively large sample size and population heterogeneity brings opportunity for estimating heterogeneous treatment effects (HTE), also known as conditional average treatment effects (CATE), which quantify how treatment benefits and risks vary across patients. Accurate estimation of HTE is essential for precision medicine, enabling identification of patients most likely to benefit, or be harmed, by a given treatment. These insights could support evidence-based regulatory decisions, improve clinical guideline development, and enhance clinical decision support by tailoring treatment recommendations to patient subgroups with differential risk–benefit profiles [5].

Recent methodological advances for HTE estimation include meta-learners such as the S-, T-, and X-learners[6], tree-based approaches such as causal forests[7, 8], and survival-specific extensions including causal survival forests [9]. However, most existing approaches have been developed and demonstrated on datasets with moderate-dimensional covariates, typically around dozens to lower hundreds of covariates, which are often selected due to their potential role as confounders based on domain knowledge, existing literature, or associations in the data (e.g., high-dimensional propensity score [10]). This scale is not compatible with modern real-world health data like EHRs and claims, which typically include tens of thousands unique concepts across concept domains (e.g., diagnoses, medications, procedures, measurements) for a comparative treatment effect estimation study [11, 12, 13], and the true confounders are not directly observable and cannot be reliably specified a priori.

To avoid missing important confounders in any form of confounder selection, some real-world studies apply large-scale propensity score (LSPS), a disease-agnostic confounding adjustment technique that adjusts for (nearly) all pretreatment covariates from a real-world health database [14, 15]. LSPS was able to adjust for confounders that are not explicitly included in the adjustment set, suggesting that adjusting for large amount of observed EHR features can better pinpoint indirectly observed (i.e., latent) confounders, reducing the risk of unmeasured confounding [15]. Similar observations were made in the literature of proxy variables for causal inference, where researchers suggested that including many noisy proxy variables can lead to better confounding control, because proxy variables can lead to better capture of the latent confounder [16].

However, directly incorporating all observed covariates does not necessarily yield the most efficient estimator, as the confounding information relevant for adjustment likely lies in a much lower-dimensional latent space. For example, type 2 diabetes with poor glycemic control, a (latent) health condition that is critical to adjust for in cardiovascular studies, may be encoded by hundreds of observed covariates, including numerous diagnosis codes for type 2 diabetes, medication codes for anti-glycemic drugs, and laboratory codes for glucose-related laboratory tests. This observation suggests that although EHR data are represented by extremely high-dimensional observed features, the underlying clinical states that drive confounding may lie in a much lower-dimensional space, creating an opportunity to improve treatment effect estimation through representation learning.

Representation learning offers a principled alternative to manual feature selection for representing high-dimensional data in a low-dimensional space. Variational autoencoder (VAE) [17, 18] is one of the most widely used representing learning model. VAE has been adapted for causal inference[19, 20] and has further been extended to learn disentangled latent factors such as Treatment Effect Disentaglement Variational Autoencoder (TEDVAE) [21]. Approaches like TEDVAE learn low-dimensional representations that separate confounders, instruments, and outcome-dependent factors. This causal disentanglement can be beneficial because prior work has shown that including instrumental variables can increase variance of effect estimates and unmask bias when there is unmeasured confounding [22, 23, 24, 25, 26]. However, existing VAE-based approaches for treatment effect estimation focus primarily on continuous or binary outcomes and do not explicitly model time-to-event outcomes. Survival-focused VAE extensions [27, 28] emphasize predictive performance rather than counterfactual causal estimation.

In this study, we propose SurvTEDVAE, a disentangled variational autoencoder tailored for survival outcomes. SurvTEDVAE extends TEDVAE by incorporating a survival likelihood into the generative framework, enabling joint modeling of high-dimensional covariates, treatment assignment, and time-to-event outcomes. The model learns three disjoint latent spaces corresponding to instrumental factors, confounders, and outcome-dependent risk factors, and uses the confounders and outcome-dependent risk factors for personalized treatment effect estimation. We demonstrate that SurvTEDVAE achieved lower estimation error on real-world EHR with high-dimensional covariates.

## 2 Methods

### 2.1 Problem Setup

#### Notation

We formulate the problem of conditional average treatment effect (CATE) estimation under the potential outcome framework for causal inference [29]. Consider an observational dataset containing *N* patients indexed by *i* ∈ {1, 2, …, *N*}. For patient *i*, let *X*_*i*_ ∈ ℝ^*p*^ denote a *p*-dimensional vector of observed covariates and *T*_*i*_ ∈ {0, 1} denote the treatment assignment. Let *Y*_*i*_ denote the potential survival time and *C*_*i*_ denote the censoring time. The observed follow-up time is defined as *U*_*i*_ = min(*Y*_*i*_, *C*_*i*_) and the event indicator is Δ_*i*_ = *I*(*Y*_*i*_ ≤ *C*_*i*_) [30]. The observed dataset is therefore 𝒟 = {(*U*_*i*_, Δ_*i*_, *X*_*i*_, *T*_*i*_) : *i* = 1, …, *N*}.

#### Estimand

To define causal effects, we introduce counterfactual survival outcomes. Let *Y*_*i*_(1) and *Y*_*i*_(0) denote the potential survival times under treatment and control, respectively. We define the conditional average treatment effect (CATE) for survival outcomes using restricted mean survival time (RMST):

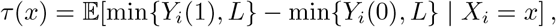

where *L* is the maximum follow-up time in the study. The CATE quantifies how treatment effects vary across patient subpopulations and provides a basis for personalized treatment recommendations in time-to-event settings.

#### Assumptions for Identification

To identify causal effects from observational data with censoring, we assume the following standard conditions.

Assumption 1 (Stable Unit Treatment Value Assumption (SUTVA)) 1) Consistency: *Y*_*i*_ = *Y*_*i*_(*T*_*i*_); 2) No interference between treatment groups.

Assumption 2 (No Unmeasured Confounding) (*Y*_*i*_(1), *Y*_*i*_(0)) ⊥ *T*_*i*_ | *X*_*i*_.

Assumption 3 (Overlap) 0 <*P* (*T*_*i*_ = 1 | *X*_*i*_) < 1.

Assumption 4 (Ignorable Censoring) *Y*_*i*_(*t*) ⊥ *C*_*i*_ | (*X*_*i*_, *T*_*i*_) for *t* ∈ {0, 1}.

Assumption 5 (Positivity of Censoring) *P* (*C*_*i*_ ≥ *u* | *X*_*i*_, *T*_*i*_) *>* 0 for all *u* ≤ *L*.

Because SurvTEDVAE performs confounding adjustment using latent representations rather than the original observed covariates, we additionally assume that the learned latent variables capture the relevant confounding information.

Assumption 6 (Latent Sufficiency) Let *Z*_*c*_ = *f*_*c*_(*X*) and *Z*_*y*_ = *f*_*y*_(*X*) denote latent representations learned from *X*. We assume

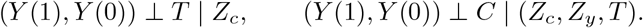

Intuitively, *Z*_*c*_ captures confounding factors that influence both treatment and outcome, while *Z*_*y*_ captures prognostic factors that affect the survival outcome but not treatment assignment.

Theorem 1 (Identification of CATE VIA *Z*_*c*_) Under Assumptions 1–6, the conditional average treatment effect is identifiable by conditioning on the confounder representation *Z*_*c*_:

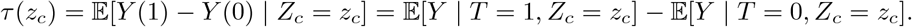

Consequently, identification also holds when conditioning on additional latent variables that include *Z*_*c*_, such as *τ* (*z*_*c*_, *z*_*t*_, *z*_*y*_), *τ* (*z*_*c*_, *z*_*t*_), or *τ* (*z*_*c*_, *z*_*y*_).

#### Error bounds for CATE estimation

While identification requires adjusting only for confounders *Z*_*c*_, incorporating additional prognostic information can improve estimation accuracy. We evaluate estimation accuracy using the precision in estimation of heterogeneous effects (PEHE), a commonly used metric for CATE estimation.

Theorem 2 (Bound for PEHE) Consider a representation-based CATE estimator that conditions on representation *Z*. Under the generalization bound of Johansson et al [31], the PEHE satisfies

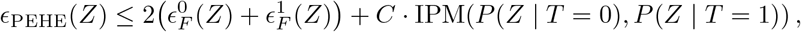

where 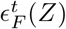 denotes the factual outcome prediction error in treatment arm *t* using representation *Z*.

If *Z* = (*Z*_*c*_, *Z*_*y*_) includes additional prognostic information beyond *Z*_*c*_, then

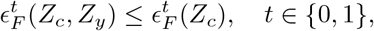

and therefore *ϵ*_PEHE_(*Z*_*c*_, *Z*_*y*_) admits a tighter upper bound than *ϵ*_PEHE_(*Z*_*c*_).

Proofs for both theorems are provided in the Appendix A.1 and A.2.

### 2.2 SurvTEDVAE: Model for estimating CATE

The proposed method, SurvTEDVAE, is a deep learning method for estimating CATE from high-dimensional datasets. SurvTEDVAE has two major components. The first component is a disentangled representation learning component: it trains a multi-task supervised variational autoencoder (VAE) to learn disentangled latent representations **Z**_**t**_, **Z**_**c**_, and **Z**_**y**_. SurvTEDVAE extends the original TEDVAE framework, which supports only continuous or binary outcomes, to accommodate time-to-event outcomes. Second, a subset of the learned representations, specifically **Z**_**c**_ and **Z**_**y**_, is incorporated into doubly robust CATE estimators (e.g., R-learner, causal survival forests, or targeted maximum likelihood estimation) to estimate treatment effects within subpopulations.

The model consists of three encoder networks that infer latent representations: 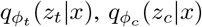, and 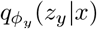. Standard Gaussian priors are assumed for all latent variables. The generative models for treatment and covariates are

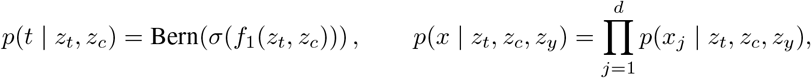

where *f*_1_ is a neural network and *σ*(·) is the logistic function. The dimensions 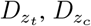, and 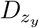 determine the size of the latent instrumental, confounding, and prognostic representations.

For time-to-event outcomes, the survival time *y* is modeled using an exponential distribution with rate parameter determined by neural networks:

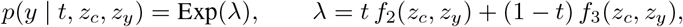

where *f*_2_ and *f*_3_ are neural networks and a Softplus activation ensures λ > 0.

The inference model uses Gaussian variational posteriors: 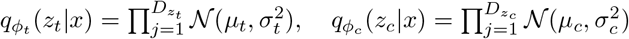 and 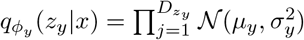. Model training maximizes an evidence lower bound (ELBO) consisting of reconstruction, treatment, outcome, and regularization terms:

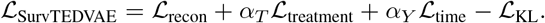

The reconstruction loss ensures that the latent variables retain information about the observed covariates. The treatment loss encourages the latent representation (*z*_*t*_, *z*_*c*_) to capture treatment assignment mechanisms. The outcome loss models the observed survival time using the exponential likelihood with censoring indicator Δ_*i*_. Finally, the KL divergence regularizes the latent variables toward their Gaussian priors. The full loss components are 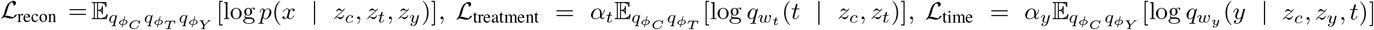, and 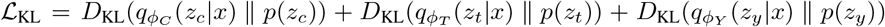. These components encourage the model to learn disentangled latent representations that reconstruct covariates, treatment, and survival outcomes, enabling counterfactual outcome generation for causal inference. The implementation of SurvT-EDVAE is available at https://github.com/wpowell31/SurvTEDVAE/.

### 2.3 Data sources

We evaluated models on two datasets: 1) ACTG-Synthetic dataset and 2) EHR-HTN dataset. The ACTG-Synthetic dataset was introduced by Chapfuwa et al. [32], which is derived from the AIDS Clinical Trials Group (ACTG) Study 175, a randomized clinical trial that evaluated treatments in adults infected with human immunodeficiency virus (HIV) [33]. The dataset preserves the real-world covariate distribution of the original ACTG trial while simulating a non-random treatment assignment, time-to-event outcomes generated from a Gompertz-Cox model, and censoring times drawn from an accelerated failure time (AFT) mechanism, following the procedure described in Chapfuwa et al. The resulting dataset includes 23 baseline covariates for 2,139 individuals.

The EHR-HTN dataset was derived from the Washington University/BJC HealthCare (WashU) EHR database, which includes data from 14 hospitals in the St. Louis metropolitan area for about 6 million patients. We followed the cohort selection criteria outlined in [11] to construct the exposure and outcome cohorts and named the dataset EHR-HTN. The dataset includes patients diagnosed with hypertension and receiving either thiazide/thiazide-like diuretics (THZ), the target exposure, or angiotensin-converting enzyme inhibitors (ACEi), the comparator exposure. The outcome is time to acute myocardial infarction. For this study, we preserve the real-world covariate distribution and simulate the treatment assignment and outcomes following procedures similar to those used in the ACTG-Synthetic dataset. The dataset includes 20,031 covariates for 45,142 patients.

### 2.4 Experiments

We compare SurvTEDVAE against existing HTE estimators on both datasets for CATE estimation.

#### Experimental design

For both the ACTG-Synthetic and EHR-HTN experiments, the data were split into training, validation, and test sets using a 60–30–10 ratio. Hyperparameters for all models were tuned using the validation set. For SurvTEDVAE, we performed grid search over the learning rate, learning rate decay, dimensionality of each latent variable cluster ({5, 10, 15, …, 50}), and number of hidden layers ({3, 4, 5}). For comparison methods, S-Learner, T-Learner, and X-Learner meta-learners were implemented following the configuration described by Xu et al. [34]. The Causal Survival Forest was implemented in `R` using the `grf` package [35].

#### Evaluation metrics

To evaluate the accuracy of CATE estimation, we used *Precision in Estimation of Heterogeneous Effect* (PEHE) [36, 37]. PEHE measures the discrepancy between the estimated and true individual treatment effects: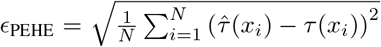, where 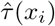 denotes the estimated CATE for individual *i, τ* (*x*_*i*_) is the ground truth CATE, and *N* is the number of individuals. PEHE captures both bias and variance in individual treatment effect estimation.

We also evaluated the population-level average treatment effect (ATE) estimation error to assess how well individual CATE estimates aggregate to the true ATE. The ATE error is defined as: 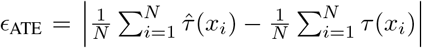 where 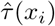 is the estimated CATE and *τ* (*x*_*i*_) is the ground truth CATE. The ATE error measures the absolute difference between the average estimated and average true treatment effects at the population level. All evaluation metrics were computed on the test set. We employed 10 Monte Carlo samples and reported the standard deviation across Monte Carlo samples.

## 3 Results

### 3.1 Results from ACTG Study

#### Main results

Table 1 presents the performance of SurvTEDVAE and comparison methods on the ACTG dataset for the 365-day follow-up horizon. SurvTEDVAE achieves the lowest PEHE (13.14 ± 0.04), outperforming competing methods including S-Learner (13.99 ± 0.13), T-Learner (14.78 ± 0.44), CSF (17.35 ± 1.04), and X-Learner (16.52 ± 1.04). The model’s PEHE performed better with a higher number of generative samples drawn during inference. SurvTEDVAE also achieves competitive ATE error (2.19 ± 0.83), comparable to CSF (2.45 ± 1.38) and X-Learner (2.35 ± 0.74), indicating that accurate individual-level CATE estimates aggregate well to the population-level treatment effect. Notably, SurvTEDVAE demonstrates the smallest variability in PEHE across repeated experiments, suggesting improved stability in treatment effect estimation.

**Table 1.** Model performance in ACTG study.

| Method | $\epsilon_{\text{PEHE}}$ | $\epsilon_{\text{ATE}}$ |
| --- | --- | --- |
| SurvTEDVAE | **$13.14 \pm 0.04$** | **$2.19 \pm 0.83$** |
| CSF | $17.35 \pm 1.04$ | $2.45 \pm 1.38$ |
| S-Learner | $13.99 \pm 0.13$ | $5.88 \pm 0.21$ |
| T-Learner | $14.78 \pm 0.44$ | $4.38 \pm 0.49$ |
| X-Learner | $16.52 \pm 1.04$ | $2.35 \pm 0.74$ |
| Neural Network | $18.13 \pm 3.24$ | $6.39 \pm 0.53$ |

**Table 2.** Model performance in EHR-HTN study.

| Method | $\epsilon_{\text{PEHE}}$ | $\epsilon_{\text{ATE}}$ |
| --- | --- | --- |
| SurvTEDVAE | **$181.11 \pm 0.04$** | **$0.67 \pm 0.21$** |
| CSF | $183.59 \pm 3.72$ | $9.50 \pm 4.31$ |
| S-Learner | $182.32 \pm 0.12$ | $0.96 \pm 0.50$ |
| T-Learner | $222.98 \pm 9.61$ | $44.14 \pm 25.56$ |
| X-Learner | $203.75 \pm 9.99$ | $33.25 \pm 19.88$ |
| Neural Network | $244.70 \pm 0.33$ | $9.24 \pm 0.75$ |

Results for the longer 730-day follow-up horizon are reported in Appendix A.3, where SurvTEDVAE similarly achieves the lowest PEHE among the evaluated methods. These findings suggest that SurvTEDVAE’s learned representations improve both individual-level and population-level treatment effect estimation compared with existing causal survival approaches.

Figure 2 provides deeper insight into SurvTEDVAE’s performance by comparing estimated and ground truth CATE distributions. SurvTEDVAE’s estimated CATE distributions align more closely with the ground truth trend (shown by the dashed line) across the full range of true CATE values. In contrast, metalearners exhibit distinct failure modes. S-Learner and T-Learner produce CATE estimates that are systematically biased toward zero, failing to capture heterogeneous treatment effects. Causal Survival Forest and X-Learner better capture the overall trend in true CATE but exhibit substantially larger variance in their estimates. All methods, including SurvTEDVAE, show elevated prediction errors when the ground truth CATE is negative, likely due to the relatively small number of samples in this region. Overall, SurvTEDVAE maintains the best bias–variance tradeoff, achieving both accurate trend estimation and low variance across the range of CATE values.

**Figure 1.**
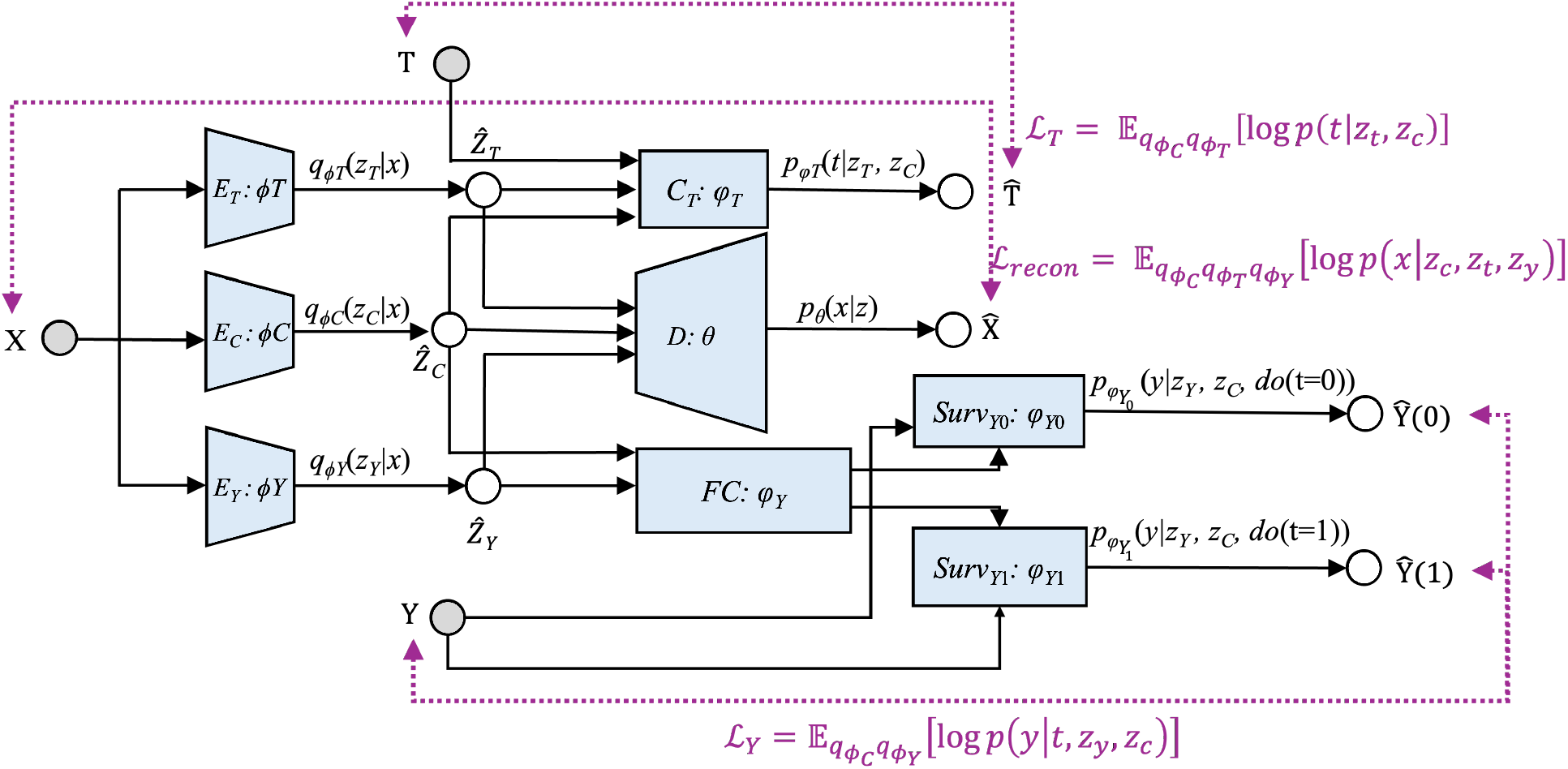
SurvTEDVAE architecture. Dotted arrows indicate components of the loss function.

**Figure 2.**
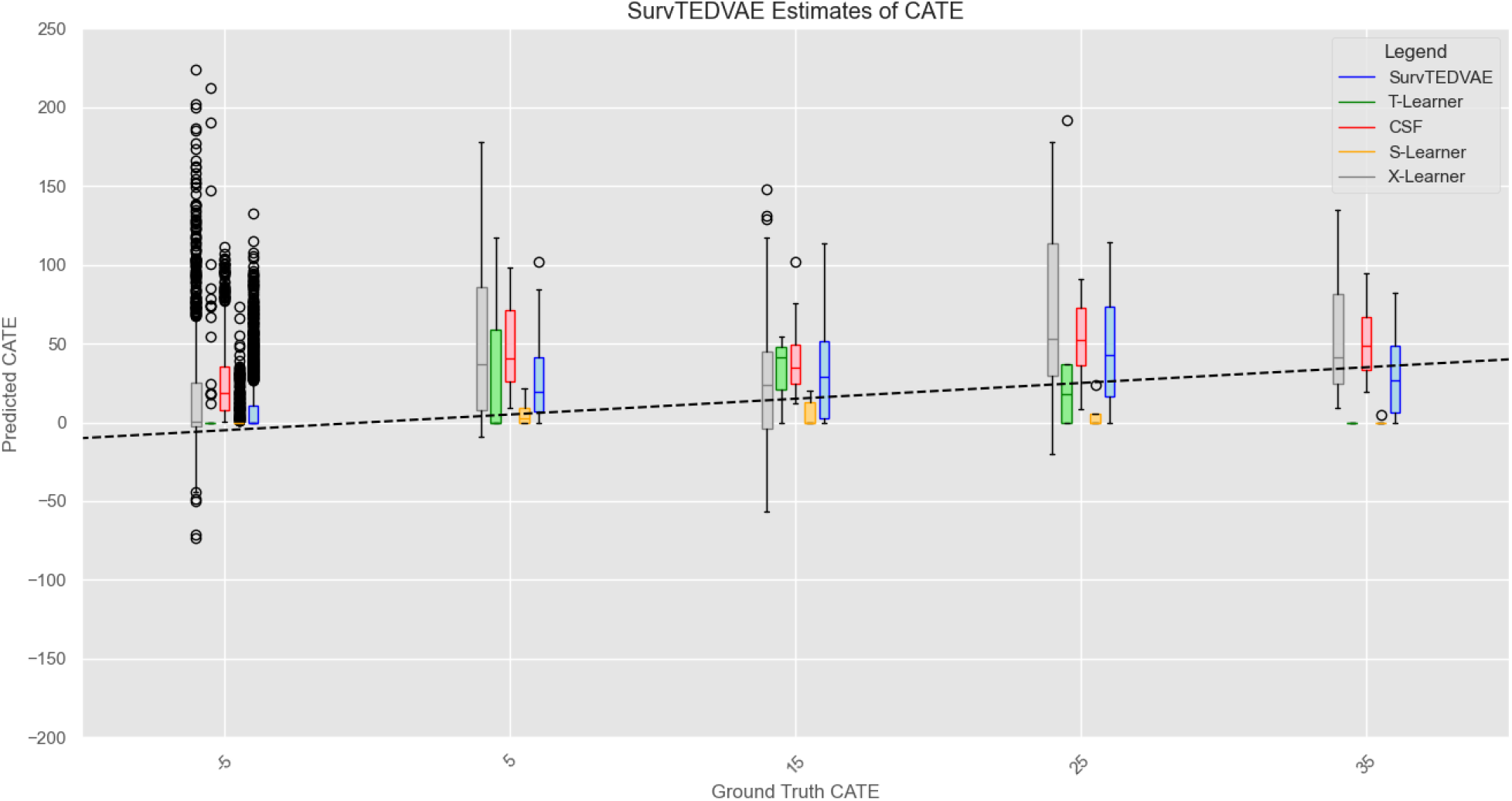
Predicted versus ground truth CATE in the ACTG study. SurvTEDVAE captures heterogeneous treatment effects with smaller errors across the range of true CATE values.

### 3.2 Results from Real-World Study

We evaluated SurvTEDVAE on the EHR-HTN dataset, which contains high-dimensional data (20,031 covariates). SurvTEDVAE achieves the lowest PEHE (181.11 ± 0.04) and ATE error (0.67 ± 0.21) among all competing methods. The tight confidence intervals around SurvTEDVAE’s estimates indicate stable performance across different data partitions. In contrast, metalearners (T-Learner, X-Learner) show substantially higher ATE errors with wide confidence intervals, suggesting unstable estimates on this high-dimensional dataset. These results demonstrate SurvTEDVAE’s ability to leverage representation learning for confounding adjustment in realistic, high-dimensional electronic health record data.

## 4 Discussion

In this study, we proposed SurvTEDVAE, a disentangled variational autoencoder designed for estimating heterogeneous treatment effects (HTE) with time-to-event outcomes. By integrating survival likelihood modeling with disentangled representation learning, SurvTEDVAE enables causal effect estimation in settings characterized by high-dimensional covariates and censoring, which are common in real-world clinical data such as EHRs. Across both a semi-synthetic benchmark dataset and a high-dimensional EHR dataset, SurvTEDVAE consistently achieved lower estimation error compared with existing meta-learning and tree-based approaches for causal survival analysis.

Our results highlight the feasibility of disease-agnostic automated dimension reduction for transforming high-dimensional EHR features into low-dimensional latent representations using deep learning models. Such dimensionality reduction can improve causal effect estimation because directly modeling extremely high-dimensional inputs could potentially introduce noise, increase estimator variance, and reduce the stability and efficiency of treatment effect estimation [38, 10, 37]. SurvTEDVAE addresses this challenge by learning disentangled latent representations that separate instrumental factors, confounders, and prognostic outcome factors. This disentanglement allows the model to focus on representations most relevant for confounding adjustment while avoiding variance inflation from irrelevant or instrumental variables, thereby improving the precision of effect estimation.

From a methodological perspective, SurvTEDVAE contributes to the growing literature on deep generative models for causal inference by extending disentangled variational autoencoder frameworks to survival outcomes with censoring. Several recent works have also applied representation learning to survival analysis. For example, the Survival Analysis Variational Autoencoder (SAVAE) extends the VAE architecture to model time-to-event data for predictive survival analysis [27]. Similarly, DAGSurv integrates causal graphical structures with variational autoencoders to improve survival prediction [28]. However, these methods are primarily designed for predictive modeling and do not address causal effect estimation. In contrast, SurvTEDVAE is designed specifically for causal inference with survival outcomes. By incorporating a survival likelihood within the generative model, SurvTEDVAE jointly models covariates, treatment assignment, and survival time while accounting for censoring. This formulation allows the learned latent representations to support downstream doubly robust estimators for heterogeneous treatment effect estimation.

Despite these promising results, several limitations should be considered. First, evaluation of individual-level treatment effects requires semi-synthetic datasets where counterfactual outcomes are available. Although such benchmarks are widely used for methodological validation in causal inference research, additional evaluations using diverse data-generating mechanisms and real-world validation studies would further strengthen evidence for the method. Second, the current implementation models survival times using an exponential distribution, which may not fully capture more complex hazard structures observed in clinical datasets. Extending the framework to more flexible parametric or nonparametric survival models, such as Weibull distributions or neural hazard functions, may further improve performance. Third, the use of deep generative models introduces computational overhead and requires careful hyperparameter tuning. Future work may explore more efficient training strategies or alternative representation learning architectures that reduce computational burden. Finally, interpretability remains a challenge for deep learning models used in biomedical research. Although SurvTEDVAE provides a structured decomposition of latent factors into instrumental, confounding, and prognostic components, additional work is needed to improve the interpretability of these latent representations and to link them to clinically meaningful features. Visualization of latent spaces and post-hoc explanation methods may help improve transparency and facilitate adoption in clinical and biomedical research settings.

In summary, SurvTEDVAE provides a new framework for heterogeneous treatment effect estimation with survival outcomes in high-dimensional observational data. By combining disentangled representation learning with causal survival modeling, the proposed method achieves improved accuracy and stability compared with existing approaches. These results suggest that deep generative representation learning may play an important role in advancing causal inference for large-scale biomedical data, particularly in applications involving electronic health records and other complex real-world data sources.

## Data Availability

The datasets generated and analyzed during this study are available from the corresponding author upon reasonable request, subject to institutional data sharing agreements.

## Appendix

### A.1 Proof of Theorem 1

Fix *z*_*c*_ and *t* ∈ {0, 1}. By latent ignorability (Assumption 6), *Y* (*t*) ⊥ *T* | *Z*_*c*_, hence 𝔼 [*Y* (*t*) | *Z*_*c*_ = *z*_*c*_] = 𝔼 [*Y* (*t*) | *T* = *t, Z*_*c*_ = *z*_*c*_]. By consistency (Assumption 1), we have 𝔼 [*Y* (*t*) | *T* = *t, Z*_*c*_ = *z*_*c*_] = 𝔼 [*Y* | *T* = *t, Z*_*c*_ = *z*_*c*_]. Combining the two equalities yields 𝔼 [*Y* (*t*) | *Z*_*c*_ = *z*_*c*_] = 𝔼 [*Y* | *T* = *t, Z*_*c*_ = *z*_*c*_] for *t* ∈ {0, 1}. Therefore,

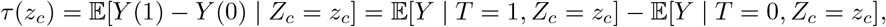

which proves identification of *τ* (*z*_*c*_). Consequently, identification also holds when conditioning on additional latent variables that include *z*_*c*_, such as *τ* (*z*_*c*_, *z*_*t*_, *z*_*y*_), *τ* (*z*_*c*_, *z*_*t*_), or *τ* (*z*_*c*_, *z*_*y*_).

### A.2 Proof of Theorem 2

The bound follows from Shalit et al [37]. The discrepancy term depends on the induced representation distribution across treatment groups and is controlled primarily by confounding structure captured by *Z*_*c*_. Adding an outcome-predictive representation *Z*_*y*_ can reduce the factual prediction errors 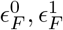 without increasing imbalance when *Z*_*y*_ is primarily prognostic. Therefore the PEHE upper bound using (*Z*_*c*_, *Z*_*y*_) is no larger than the bound using *Z*_*c*_ alone.

### A.3 Results in ACTG Study with 730-day Follow-up Period

| Metric | SurvTEDVAE | CSF | S-Learner | T-Learner | X-Learner | Neural Network |
| --- | --- | --- | --- | --- | --- | --- |
| $\epsilon_{\text{PEHE}}$ | **75.02** | 85.47 | 78.17 | 79.00 | 85.98 | 83.96 |
| $\epsilon_{\text{ATE}}$ | **11.78** | 14.54 | 37.44 | 25.96 | 11.91 | 16.85 |

## Notes

### Competing Interest Statement

The authors have declared no competing interest.

### Funding Statement

No external funding was received.

### Author Declarations

This study was approved by the Washington University Institutional Review Board (IRB) under IRB protocol 202604175.

